# Predicting preterm births from electrohysterogram recordings via deep learning

**DOI:** 10.1101/2022.12.25.22283937

**Authors:** Uri Goldsztejn, Arye Nehorai

**Affiliations:** Department of Biomedical Engineering, McKelvey School of Engineering, Washington University in St. Louis, St. Louis, MO, USA; Preston M. Green Department of Electrical and Systems Engineering, McKelvey School of Engineering, Washington University in St. Louis, St. Louis, MO, USA

## Abstract

About one in ten babies is born preterm, i.e., before completing 37 weeks of gestation, which can result in permanent neurologic deficit and is a leading cause of child mortality. Although imminent preterm labor can be detected, predicting preterm births more than one week in advance remains elusive. Here, we develop a deep learning method to predict preterm births directly from electrohysterogram (EHG) recordings of pregnant mothers without symptoms of preterm labor. We developed a prediction model, which includes a recurrent neural network, to predict preterm births using short-time Fourier transforms of EHG recordings and clinical information from two public datasets. We predicted preterm births with an area under the receiver-operating characteristic curve (AUC) of 0.80 (95% confidence interval: 0.79-0.82). Moreover, we found that the spectral patterns of the measurements were more predictive than the temporal patterns, suggesting that preterm births can be predicted from short EHG recordings in an automated process. We show that, even without symptoms of preterm labor, preterm births can be predicted for pregnant mothers around their 31st week of gestation, prompting beneficial treatments to reduce the incidence of preterm births and improve their outcomes.

## Introduction

Around 10% of all live births, about 15 million babies per year, are preterm, that is, they happen before 37 weeks of gestation are completed [1, 2]. Preterm births are a leading cause of newborn mortality [3]. Moreover, many preterm babies suffer from long-term morbidity, including permanent neurological damage [2, 4]. Because treatments can delay preterm births and improve their outcomes, identifying pregnant mothers at high risk of preterm birth is compelling, as recognized by the World Health Organization (WHO) [2, 5].

Although several methods can predict preterm births, they have limitations. Broad historical risk factors, such as previous preterm births or multiple gestations, can identify mothers at higher risk of preterm birth, but these risk factors alone are not sufficient to accurately predict which individual mothers will deliver preterm [6, 7, 8].

In clinical practice, preterm birth is usually predicted by measuring cervical length or the concentration of cervico-vaginal fibronectin alpha [8]. In mothers with symptoms of preterm labor, these minimally invasive tests can predict births that will occur within one week [9, 10]. Moreover, the combination of these tests has been reported to produce more accurate results than each method separately and could be used to predict preterm births in symptomatic mothers within two weeks of testing [11]. These measurements are helpful because they inform physicians and guide treatments to reduce the risk of preterm labor and to improve its outcomes. However, these measurements are not cost-effective screening tools for the general population of pregnant mothers because they have low predictive values among mothers at low risk for preterm labor, such as nulliparous women with singleton pregnancies [8, 12].

Home uterine activity monitors (HUAMs) were developed to measure uterine contractions and predict preterm births. The first such devices were based on tocodynamometer recordings, which measure the pressure changes associated with uterine contractions [13]. Unfortunately, these devices could not predict preterm births, and current clinical guidelines discourage their use for this purpose [8, 13, 14].

More recently, electrohysterogram (EHG) recordings have been proposed to predict preterm births [15, 16]. EHG recordings use abdominal electrodes to measure the electrical activity associated with uterine contractions, and they can be recorded with portable devices equipped with algorithms to monitor uterine contractions [15, 17]. A variety of algorithms have been developed for predicting preterm births from various features derived from EHG measurements [15, 16, 18]. These features are generally calculated from uterine contraction intervals, either manually selected or identified using dedicated algorithms [16, 19, 20]. These intervals can also be identified with the aid of simultaneous tocodynamometer recordings [17, 19].

To the best of our knowledge, EHG measurements have not yet been shown to predict preterm births in asymptomatic mothers with a performance comparable to the clinical standards, i.e., measurements of cervical length or fibronectin alpha in mothers with symptoms of preterm labor. Although many researchers have reported nearly perfect predictions of preterm births based on EHG measurements from the “Term-Preterm EHG Database,” meticulous analysis revealed that these results were overoptimistic and resulted from data leakage [21, 22]. Namely, these works inadvertently introduced strong correlations between the data used to train the prediction models and the data used to test the performance of these models, as shown by Vandewiele et al. [16, 21, 22]. This problem was caused by inappropriate attempts to improve the models’ performance by balancing the number of term and preterm samples used to develop these models. After Vandewiele et al. corrected this problem, these models were no longer able to predict preterm births accurately [21, 22]. Additional works with sound methodology suggest that some features derived from EHG measurements can be used to distinguish between recordings of mothers who eventually delivered at term from those who delivered preterm [23, 24, 25, 26]. However, none of these works could predict preterm births with clinically useful accuracy. More recently, Xu et al. and Lou et al. developed methods for predicting preterm births avoiding data leakage [27, 28]. Although Xu et al. and Lou et al. achieved high classification performances on test sets including real and synthetic measurements, the performances of their approaches on test sets including only real measurements are not reported. Moreover, Fischer et al. used an end-to-end deep learning model to predict preterm births from EHG measurements without artificially increasing the number of preterm samples to avoid possible data leakage [29] and achieved a moderate accuracy.

Here, we present an end-to-end deep learning model that predicts preterm births directly from EHG measurements, without handcrafted features. Therefore, our model is not sensitive to varying implementations of specific features or to how uterine contractions are segmented. We developed our work using EHG measurements and supplementary clinical information from two public databases. Importantly, we developed our model with care to avoid data leakage. Using our model, we could predict preterm births in mothers without symptoms of preterm labor at around their 31st week of gestation, thus validating the observations of in [19]. The predictive accuracy was close to that achieved by using cervical length and fibronectin alpha measurements to predict preterm labors in mothers with symptoms of preterm labor and within one week of delivery. Moreover, by investigating the measurement components that contribute to the predictions of our model, we showed that it is possible to predict preterm births using short recording times, thus facilitating clinical adoption and at-home implementation of EHG measurements. This finding is aligned with the observations of Jager et al., who showed that preterm births can be predicted from short contractile or non-contractile intervals of EHG measurements with similar accuracy as when using 30-minute long recordings [19]. Our work and results encourage using EHG measurements and deep learning for predicting preterm births in real-world scenarios. Their successful employment could help reduce newborn morbidity and mortality, especially in populations with limited access to healthcare, who suffer more from preterm birth [2].

## Materials and methods

### Study participants

In developing our work, we used two datasets in the Physionet repository, aggregating data from the “Term-Preterm EHG Database” (TPEHG DB) [23] and from the “Term-Preterm ElectroHysteroGram DataSet with Tocogram” (TPEHGT DS) [19]. These datasets contain 30-minute-long bipolar EHG measurements and clinical information about pregnant mothers recorded during regular pregnancy checkups. Both datasets were acquired at the University Medical Centre Ljubljana, using the same recording protocol and device. The TPEHG DB consists of 300 records, each obtained from a different mother at either around the 22nd or the 32nd week of gestation. Additionally, the TPEHGT DS contains 26 records from 18 different mothers, obtained around the 30th week of gestation. Half of the samples in the TPEHGT DS correspond to mothers who eventually delivered preterm, while the other 13 records correspond to term deliveries. When compiling these datasets, the datasets’ authors excluded the mothers whose labors were induced or whose deliveries were performed using a Cesarean section [19, 23].

We included the records from both datasets obtained after the 28th week of gestation. Since each record in the TPEHG DB was obtained from a different mother, we included all the records from this database that were obtained after the 28th week of gestation. On the other hand, when there were multiple records for the same mother in the TPEHGT DS, we included only the latest record during the pregnancy, provided that the record was made after the 28th week of gestation. We identified the records in the TPEHGT DS that corresponded to a particular mother by comparing the clinical information. By using a single record per mother, we prevented our models from learning features that characterize mothers rather than features that are predictive of pregnancy outcomes. Overall, we used 151 records from different mothers. Among these mothers, 18.5% delivered preterm. We detail the clinical information of these mothers in Table 1.

**Table 1:** Clinical information from the records included in our work.

|  | Distribution | Predictive power | No. of missing entries |
| --- | --- | --- | --- |
| Number of participants | 151 | - | - |
| Number of preterm births (%) | 28 (18.5) | - | - |
| **Continuous predictors<sup>a</sup>** | **Median (IQR)** | **AUC (95% CI)** |  |
| Age [years] | 28 (26-32) | 0.51 (0.28-0.63) | 23 |
| Gestational age at recording [weeks/days] | 31/1 (30/6-31/6) | N/A | 0 |
| Weight [Kg] | 72 (66-76.8) | 0.54 (0.40-0.67) | 28 |
| **Binary predictors<sup>b</sup>** | **No. of positives (%)** | **Risk ratio (95% CI)** |  |
| Parous | 47 (31.1) | 1.86 (1.16-2.99) | 0 |
| Previous abortions | 21 (13.9) | 1.03 (0.38-2.83) | 0 |
| Hypertension | 0 (0.0) | N/A | 16 |
| Diabetes | 1 (0.7) | N/A | 16 |
| Placental position | 63 (51.6) | 1.50 (1.07-2.11) | 29 |
| Bleeding in 1st trimester | 12 (8.9) | 0.26 (0.02-4.27) | 16 |
| Bleeding in 2nd trimester | 3 (2.2) | 14.50 (1.24-169.16) | 16 |
| Funneling | 5 (3.7) | 1.63 (0.19-13.73) | 16 |
| Smoker | 9 (6.0) | 2.20 (0.58-8.25) | 0 |

<sup>a</sup> We characterized the data distributions using the median values and interquartile ranges (IQR). We assessed the predictive power of the continuous predictors using the area under the receiver-operating characteristic curve (AUC) and calculated the 95% confidence interval (CI) using the logit method. <sup>b</sup> For the binary predictors, we show the number of positive samples in the dataset and the percent of positive samples in the dataset (%). We assessed the predictive power, with 95% CI, of these predictors through the risk ratio test.

Additionally, we illustrate the distribution of gestational ages of the mothers included at the times of recording and at birth in Supplementary Fig. 1 and Supplementary Fig. 2.

### Prediction models

We developed classification and regression models to predict a term or a preterm birth. The classification models were trained specifically to predict categorical outcomes, i.e., delivery at term or preterm. Pursuing a different approach, we trained the regression models to predict the gestational age at delivery, labeling predictions higher than 37 completed weeks, or 259 days, as term, and those below 37 weeks as preterm. In developing the classification and regression models, we used clinical information alone, EHG measurements alone, and clinical information combined with EHG measurements. These prediction models, developed using MATLAB 2020a, are detailed in the next subsections and summarized in a block diagram in Fig. 1.

**Figure 1:**
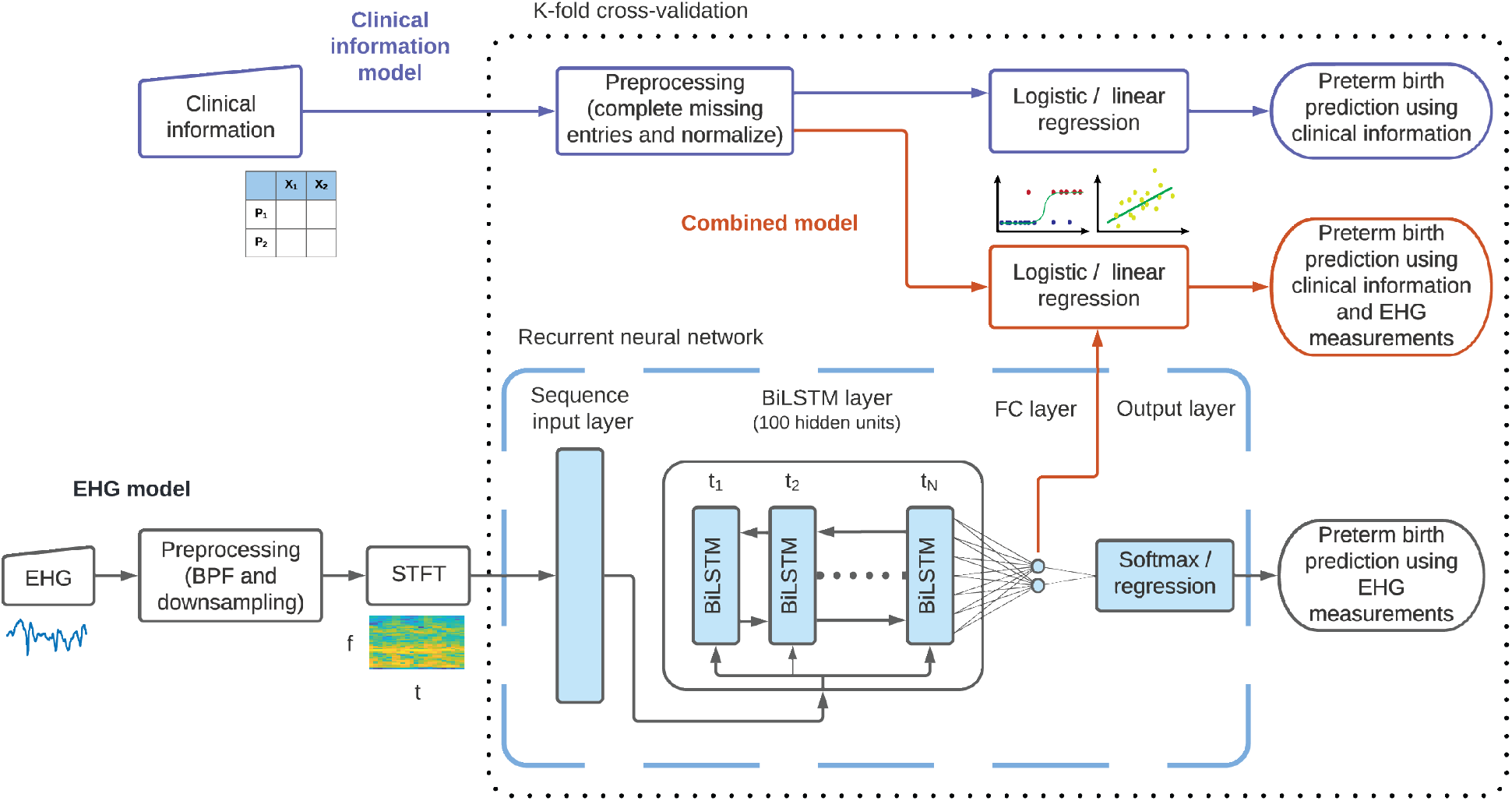
Block diagram of the three classification and regression models developed in this work. The details of these models are provided in Methods. The clinical information model is illustrated in the upper part of the diagram, using shapes with blue outlines. This model uses clinical information, in tabular format, to predict preterm births by using logistic or linear regression, represented as a block with a blue outline and schematic illustrations below it. Preprocessing the clinical information consists of completing missing entries and normalizing the predictors, as described in Methods. The EHG model is illustrated in the lower part of the diagram, using shapes with black outlines. This model uses EHG measurements, represented by an input block with a schematic illustration below it, that are first preprocessed. This preprocessing step includes bandpass filtering (BPF) and downsampling. The preprocessed measurements are used to compute STFTs, illustrated by a block and a schematic representation, that are used as input to the RNN. This network is composed of an input layer, a BiLSTM layer, a fully connected (FC) layer, and an output layer, which are illustrated using light blue shapes with black outlines and enclosed within a dashed light blue outline. The combined model uses clinical information and EHG measurements to predict preterm births and is illustrated in the middle part of the diagram using shapes with red outlines. The dotted black outline represents the cross-validation technique employed, indicating that the operations within are applied separately for each data partition, whereas the operations outside are applied to all the data, independent of the data partition.

### Clinical information models

First, using only the clinical information of the records, we predicted whether each mother delivered preterm or at term. We used most of the predictors shown in Table 1, namely maternal age, gestational age at the time of the recording, weight, whether the mothers had given birth previously (parous), had aborted pregnancies previously, had reported vaginal bleeding in the first trimester, had reported vaginal bleeding in the second trimester, or were smokers. We excluded diagnoses of hypertension and diabetes because these diagnoses are mostly absent in this dataset. We also excluded diagnoses of funnelling because they are made through transvaginal sonography and because, in this dataset, these diagnoses have a low predictive power [8]. Similar to funnelling, we excluded the variable in the dataset indicating the placental position, which takes the values “front” (considered as the positive value in Table 1) and “end.” We completed the missing entries for each variable in the training and testing datasets using the mode of that variable in the training set. To prevent data leakage, rather than using the modes of the entire dataset, we used the modes of the samples in the training set to complete missing entries in both the training and testing sets. Therefore, our training data does not contain any information from the test set and when making predictions, our model uses only information from the training set to both complete missing entries and make predictions. In other words, our model makes predictions on each sample of the testing dataset using only information from the training set.

Next, we trained a logistic regression to predict whether deliveries were preterm and a linear regression model to predict the gestational age at birth. These models are represented using a block with a blue outline on the upper part of Fig. 1. In the logistic regression model, we discarded the redundant predictors, using lasso regularization. We regularized only the classification model, and not the regression model, because we observed that the lasso regularization improved the performance of the logistic regression model slightly but marginally worsened the performance of the linear regression model. Since we regularized the logistic regression model, we also normalized the predictors in this model to prevent the regularization term from penalizing the model parameters based on the scale of the predictors. Again, to prevent data leakage, we normalized both the training and testing sets using the means and standard deviations of the samples in the training set, thus avoiding revealing information from the test set to the training set.

The operation to complete missing entries described above, together with the operation to normalize the input data, comprise the preprocessing step for the clinical information. This preprocessing step is represented in Fig. 1 as a block that is executed once for each partitioning of the data into training and testing datasets, as described below.

### EHG measurements models

Then, using only the EHG measurements, we predicted whether the mothers delivered preterm or at term. We used solely the first signal (s1) in the databases. This signal measures the electric potential difference between two electrodes aligned horizontally on the abdomen, 3.5 cm above the navel, and separated by seven cm.

We preprocessed all the EHG measurements to improve the data quality. We removed the first minute of the recordings to remove transient effects. Next, we filtered the measurements to remove baseline wander and high frequency noise. Specifically, we filtered the recordings using a fourth-order, Butterworth bandpass filter with zero-phase and cutoff frequencies of 0.05 Hz and 4 Hz. Although most uterine activity is concentrated between 0.05 Hz and 0.7 Hz, we included a higher frequency range because higher frequency components have been shown to be predictive of preterm birth [19, 30]. Finally, we downsampled the measurements to 10 Hz to improve computational speed without losing information. These preprocessing operations are represented using a block with a black outline at the bottom of Fig. 1.

Next, as illustrated in the bottom part of Fig. 1, we transformed the preprocessed EHG measurements to the time-frequency domain, using the short-time Fourier transform (STFT). We used the STFT following the positive results previously reported using this transformation for predicting preterm births from EHG measurements [19, 27]. The STFT usefully represents how the spectral components of the measurements change over time by constructing a matrix where each column corresponds to a sliding time interval and contains the estimated spectral content of the measurements during the corresponding time interval. This transformation is helpful in analyzing non-stationary processes, such as the contractile activity during the recordings. We estimated the STFT using Hamming windows of 60 s that were slid using a 75% overlap. We chose this configuration since uterine contractions usually last around one minute and because this configuration resulted in satisfactory temporal and spectral resolutions based on visual inspection [31].

We predicted the pregnancies’ outcomes from EHG recordings using a deep neural network, rather than using handcrafted features, because neural networks automatically learn the most informative features from the data [32, 33]. Given the limited success of various methods designed to predict preterm births from the EHG measurements in the TPEHG DB using handcrafted features, Vandewiele et al. suggested using deep learning to achieve better results [22].

In agreement with Vandewiele et al., we used a deep recurrent neural network (RNN) to predict the pregnancies’ outcomes from EHG measurements, developing a dedicated network architecture for this task. This RNN uses the training set, consisting of data samples labeled with their respective pregnancy outcome, to learn features from the input data that predict the pregnancies’ outcomes. The RNN consists of a series of layers that are trained to learn multiple abstractions of the data that are helpful in relating the input data to the predictions [32]. The first layer in our network is a sequence input layer that rearranges the matrices of STFTs so that the columns of the STFT matrices, which capture the spectral content of the measurements during the sliding time intervals, become a set of features for the corresponding time step in the RNN. This input layer feeds into a series of bidirectional long short-term memory (BiLSTM) cells with 100 hidden states. The BiLSTM cells are able to learn patterns from sequential data: in our case, these cells are intended to learn patterns from the spectral changes of the EHG measurements over time. Similar network architectures, using long short-term memory (LSTM) and BiLSTM cells, have been used to successfully learn informative data representations from STFTs in other applications [34, 35].

Next, using a similar approach as Zhu et al., we connected the last BiLSTM cell to a fully connected layer consisting of two neurons, and finally we connected the fully connected layer to an output layer [34]. The fully connected layer encodes the data abstraction inferred by the BiLSTM cells into a pair of scalar values, which are then used by the output layer to make a prediction. In the classification model, this pair of scalar values scores the association of each EHG recording to the preterm and term categories. This architecture is illustrated in Fig. 1.

We used two different output layers, depending on whether we intended to predict the categorical outcome of the pregnancy or to predict the gestational age at birth. For the classification problem, we used a softmax output layer and trained the network using a weighted cross-entropy loss function that penalized errors in the preterm birth predictions more. We determined the weights of the loss function based on the relative frequency of each class in the training set, a strategy that addresses the class-imbalance problem of predicting preterm births. Namely, because term labors are more frequent in the general population and in the database, classification models trained on these data are naturally biased towards predicting term labors and may learn to predict term labors for every input. This loss function is given by:

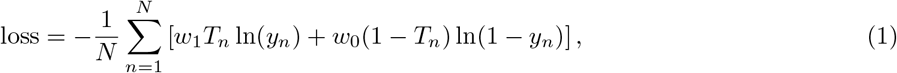

where *N* is the number of samples in each training batch, *w*_*i*_ is the penalization weight of each class, *T*_*n*_ = {0, 1} is the label of sample *n*, and *y*_*n*_ is the output score of the sample *n*. We set the penalization weight for class *i* to be:

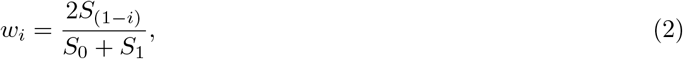

where *S*_*i*_ is the number of samples from class *i* in the training set, as suggested in [36].

For the regression problem, we used a regression layer as the output of the network. This layer implements a mean square error (MSE) loss function to train the network. Since the regression models are trained on a continuous output, i.e., the gestational age at birth, these models are less sensitive to the class imbalance problem. The classification and regression output layers are represented by a single blue block with a black outline at the bottom of Fig. 1.

In developing our prediction models using EHG measurements, we evaluated alternative model designs based on a single run of a five-fold cross-validation. We evaluated alternative time-frequency representations, namely wavelet transforms and the empirical mode decomposition, as described in [37, 38]. Additionally, we tested other neural network architectures, namely using long short-term memory (LSTM) cells and convolutional neural networks (CNN), with varying network parameters, such as the numbers of layers and the number of LSTM cells. Here, we report the model that produced the best prediction results.

We also fine-tuned the learning parameters based on a single run of a five-fold cross-validation. Namely, we selected an appropriate mini-batch size, number of training iterations, learning rate, and regularization hyperparameter.

### Combined models

We developed both a classification and a regression model that combine clinical information with EHG measurements to predict pregnancies’ outcomes. We first trained the network described in the previous subsection. Then, we extracted the activation values of the fully connected layer and concatenated these values with the clinical information. Next, we used the combined data to train the logistic regression model to predict the outcome of the pregnancy, and the linear regression model to predict the gestational age at delivery. We implemented these logistic and linear regression models as described before. The difference between these models and those used for predictions based only on clinical information is that, in this case, the data vectors included the activations of the fully connected layers in addition to the clinical information. The stages of these classification and regression models, which combine clinical information and EHG measurements, are illustrated in the middle part of Fig. 1.

### Cross-validation

We evaluated the performance of our models using a stratified five-fold cross-validation. We partitioned the data into a training set, containing 80% of the data, and a test set, containing the remaining 20% of the data, so that both the training and testing sets included the same proportion of preterm samples. We used the training set to train our models and the testing set to evaluate the models’ performance. We repeated this process five times, each time using a different set of samples for the training and testing set, so that all the samples were used for testing throughout the five runs. This cross-validation routine is indicated in Fig. 1 by a dotted black outline. This outline symbolizes that the operations represented within are applied separately for each partition of the data, whereas the operations represented outside the outline are applied once for all the data.

### Statistical analysis

To evaluate the performance of the prediction models with confidence intervals, we repeated the cross-validation routine 20 times, as recommended in [39]. Each time, we used a different random partition of the data. By repeating the cross-validation routine with various random partitions, we prevented our models from possibly producing over-optimistic results due to fitting of the training hyperparameters and model specifications to a specific cross-validation partition. We then calculated the mean and 95% confidence interval (CI) of the performance statistics, assuming that the performance statistics had Gaussian distributions with unknown means and variances.

## Results

### Performance of the prediction models

First, we attempted to predict preterm births by using only the clinical information, which supplements the EHG measurements and is described in Table 1. We developed two models: a logistic regression model to determine whether a pregnancy would result in a preterm birth, and a linear regression model to predict the gestational age at delivery, as detailed in Methods. When using the regression model, we predicted that a birth would be at term if the estimated gestational age at delivery was more than 37 complete weeks, or 259 days. The classification model predicted preterm births with an area under the receiver-operating characteristic curve (AUC) of 0.56 (95% CI: 0.54-0.59), whereas the regression model predicted preterm births with an AUC of 0.57 (95% CI: 0.54-0.59). The accuracy of clinical information alone in predicting preterm births was little more than that of a random guess.

Next, we examined whether EHG measurements could be used to predict preterm births using end-to-end deep-learning models, directly from EHG measurements and without requiring handcrafted features. Specifically, we trained a recurrent neural network to predict whether the pregnant mothers would deliver preterm and to predict their gestational ages at delivery, as described in Methods. This network’s predictions surpassed those of the clinical information models. The classification model trained on EHG measurements was able to predict preterm births with an AUC of 0.75 (95% CI: 0.74-0.77), whereas the regression model predicted preterm births with an AUC of 0.71 (95% CI: 0.68-0.73).

We also developed models to predict preterm births based on clinical information combined with EHG measurements, as described in Methods. We hypothesized that integrating the clinical information and the EHG measurements would yield more accurate prediction models, because the models trained independently on clinical information alone and EHG measurements alone could predict preterm births better than random guessing. Moreover, the clinical information and the EHG measurements provide complementary information about the pregnancy. Consistent with our hypothesis, the prediction models trained on both clinical information and EHG measurements slightly outperformed the models trained on clinical information alone and on EHG measurements alone. Our classification model predicted preterm births with an AUC of 0.80 (95% CI: 0.79-0.82), and the regression model predicted preterm births with an AUC of 0.75 (95% CI: 0.73-0.77).

To better evaluate the performance of our prediction models, we estimated a performance bound on this classification problem. In our work, as well as in the obstetrics literature and clinical practice, births are considered preterm if the mother delivers the fetus before completing 37 weeks of gestation. However, the gestational age of the mother has an uncertainty that depends on the method used to estimate it. Generally, gestational age is estimated based on a first trimester ultrasound examination or on the timing of the last menstrual period (LMP) [40]. When the gestational age is estimated based on early ultrasound examination, the estimate has a standard deviation of about five days, whereas estimates based on the LMP have standard deviations of about seven days [41]. Notably, the incidence of preterm births depends on the method used to estimate the gestational age [42].

This estimation error translates into uncertainty in the ground truth labels and limits the possible performance of classification algorithms. We estimated the upper bound of the AUC due to this limitation by measuring the AUC obtained when predicting the gestational age at delivery using a noisy version of the true gestational ages at delivery. We corrupted the gestational ages at delivery by adding independent and identically distributed (i.i.d.) Gaussian noise with zero mean and a standard deviation of six days. After repeating this procedure 20 times to estimate the mean and 95% CI of this AUC using this approach, we found that the upper AUC bound for this classification problem is 0.97 (95% CI: 0.97-0.97).

In Fig. 2, we present the receiver-operating characteristic curves (ROC) for the classification and regression models trained on clinical information alone, EHG measurements alone, and clinical information combined with EHG measurements. We observe that the classification models outperform the regression models trained on the same data. Moreover, we notice that regardless of whether we use the classification or regression approach, the EHG-based models outperform the clinical information-based models and that the models that leverage both the clinical information and the EHG measurements achieve the best performance.

**Figure 2:**
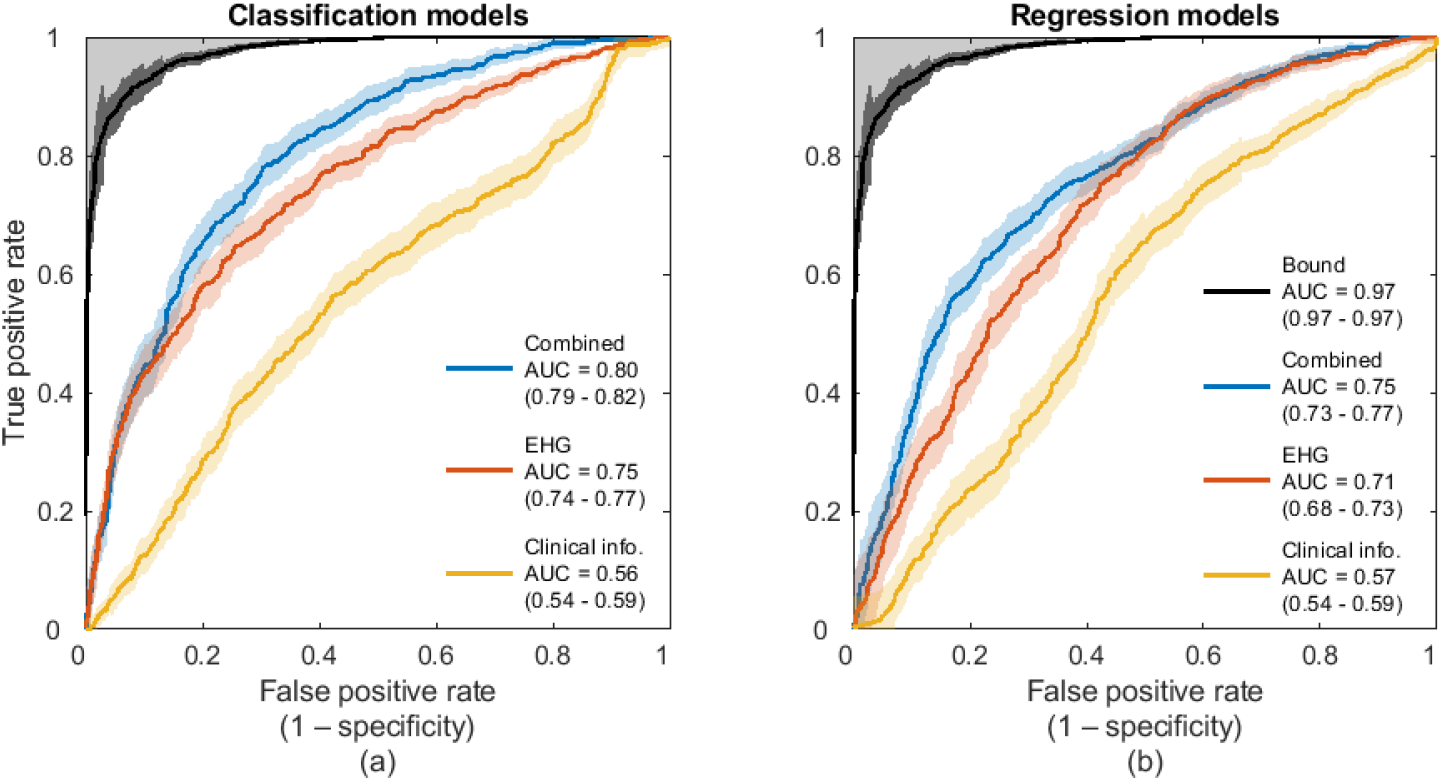
Performance of the models for predicting preterm births. (a) ROC curves for predicting preterm births using the classification models trained with clinical information alone, EHG measurements alone, and clinical information combined with EHG measurements. (b) ROC curves for the same tasks as in (a), but using the regression models instead of the classification models. (a), (b) The performance bound is shown in both panels by a black ROC curve. The greyed area delimited by this bound indicates unattainable performance due to the uncertainty in the ground truth labels. The AUCs of the models are presented with 95% CIs.

To further assess the performance of our models, we measured the sensitivity, positive predictive value (PPV), and negative predictive value (NPV) at various specificity levels, as shown in Table 2 [43]. Since the classification models systematically outperformed the regression models, we present the results only for the classification models. Since the PPV and NPV depend on the incidence of preterm births in the dataset and our dataset over-represents preterm births, we randomly removed preterm samples from each prediction fold when estimating the PPV and NPV so that the preterm birth incidence in our dataset was similar to the incidence in the TPEHG DB [43].

**Table 2:** Performance of classification models in predicting preterm birth.

| 20em | Specificity |  |  |
| --- | --- | --- | --- |
|  | 50% | 70% | 90% |
| **Sensitivity (%)** |  |  |  |
| Clinical information | 61.6 (57.8-65.4) | 41.8 (37.9-45.7) | 12.5 (9.4-15.6) |
| EHG | 81.6 (78.8-84.4) | 67.1 (63.3-71.0) | 43.0 (38.5-47.5) |
| Combined | **89.5** (87.0-92.0) | **77.3** (73.9-80.8) | **44.1** (38.8-49.4) |
| **PPV (%)** |  |  |  |
| Clinical information | 16.5 (15.4-17.6) | 20.0 (18.3-21.6) | 19.3 (15.1-23.6) |
| EHG | 21.3 (20.6-21.9) | 28.0 (26.9-29.2) | 45.9 (43.2-48.6) |
| Combined | **22.9** (22.4-23.4) | **30.9** (29.9-31.9) | **45.9** (42.6-49.3) |
| **NPV (%)** |  |  |  |
| Clinical information | 88.7 (87.5-89.9) | 88.9 (88.0-89.7) | 86.7 (86.1-87.2) |
| EHG | 94.8 (93.9-95.6) | 93.9 (93.1-94.7) | 92.0 (91.3-92.8) |
| Combined | **97.3** (96.6-98.0) | **96.1** (95.3-96.9) | **92.1** (91.2-93.0) |

All values are presented with 95% CIs. Bold fonts indicate the best performing model for each metric and for the different specificity levels. PPV, positive predictive value; NPV, negative predictive value.

In Table 2, we observe that the combined model outperforms the models trained on clinical information alone or EHG measurements alone in sensitivity, PPV, and NPV at various specificity levels. Moreover, we observe that our models have a much higher NPV than PPV, which results from the low incidence of preterm births. In other words, our predictions of term births are more reliable than our predictions of preterm births.

We verified that our model was not discriminating between the two datasets used in our work. The TPEHG DB and the TPEHGT DS datasets were acquired with the same device and following the same protocol, so we did not expect that our model would discriminate between the samples of either dataset. We confirmed that our model does not assign one label to the samples from one dataset and another label to the samples of the other dataset. Moreover, when we trained the classification models using only the TPEHG DB, we obtained similar AUCs to those obtained when we trained the models using data from both datasets.

Although our regression models could predict preterm births more accurately than random guessing, these models were not able to predict the gestational ages at delivery with a much lower MSE than the MSE obtained using the mean gestational age at delivery in the training set, i.e., the minimum MSE estimator. Although the correlation between the predicted and true gestational ages at delivery is positive, the accuracy of the predictions is low, as shown in Supplementary Fig. 3.

### Predictive components of EHG measurements

Further, we investigated how various components of the EHG measurements contribute to the preterm birth predictions by altering the STFT representations of the data. We first explored the predictive power of various frequency bands, as shown in Fig. 3a and b. We extracted four frequency bands (B0 through B3) by using only the relevant rows of the STFT for training and testing. We considered similar frequency bands as Jager et al.: in our case, B0, B1, B2, and B3, cover the frequency ranges between 0.05 Hz and 1.0 Hz, 1.0 Hz and 2.2 Hz, 2.2 Hz and 3.5 Hz, and 3.5 Hz and 5.0 Hz, respectively [19]. The only difference between our spectral partition and that proposed by Jager et al. is that, in our case, the lower frequency cutoff of B0 is Hz instead of 0.08 Hz [19]. According to Jager et al., B0 mostly contains electrical activity associated with uterine contractions, whereas the higher bands contain harmonic frequencies of uterine reverberation caused by maternal cardiac activity [19]. Notably, we observed that the models trained on higher frequency bands achieved higher AUCs, as shown in Fig. 3b.

**Figure 3:**
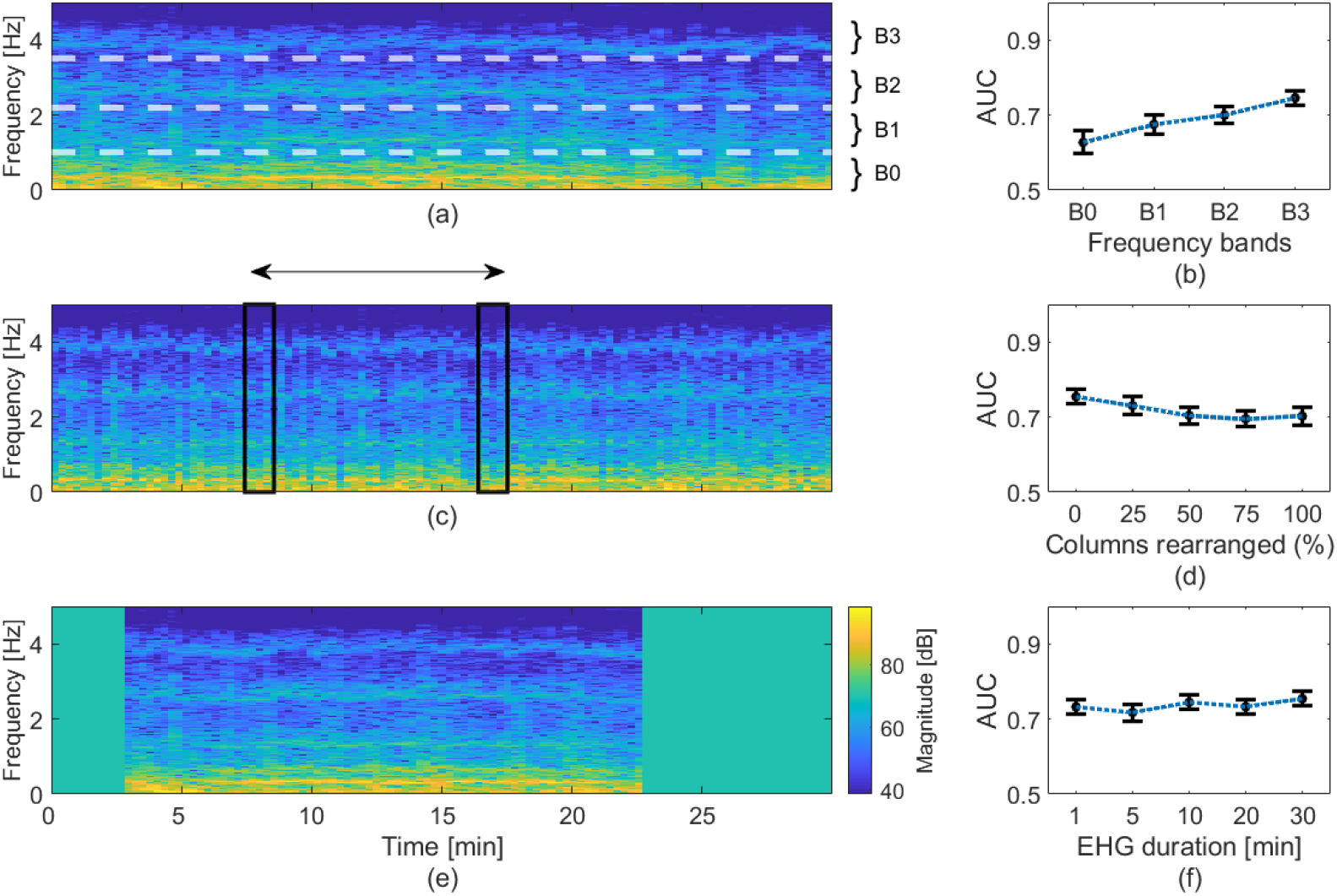
Effects of information loss on the prediction of preterm births. (a) A representative STFT of an EHG recording overlaid with the limits of the frequency bands examined. (b) AUCs obtained using the classification model trained on the various frequency bands. (c) The same STFT as in (a), but with all the columns randomly rearranged. (d) AUCs obtained using the classification model trained on STFTs with varying fractions of columns randomly rearranged. (e), The same STFT as in (a), but where ten minutes of the recording were removed. The colorbar in this panel also corresponds to panels (a) and (c). (f) AUCs obtained using the classification model trained on STFTs with varying durations. (b), (d), (f), The AUCs are presented as black dots with error bars denoting the 95% CIs.

Next, we examined how the temporal patterns of the measurements contribute to the models’ predictions. We disrupted the temporal patterns by randomly rearranging a random subset of columns of the STFTs, as illustrated in Fig. 3c. Although the AUC of the model decreased as larger fractions of columns of the STFTs were rearranged, this decline was moderate, as shown in Fig. 3d. Notably, when all the columns of the STFTs were randomly rearranged, i.e., when all the temporal patterns were disrupted, our classification model trained on disrupted EHG measurements alone was able to predict preterm births with an AUC of 0.70 (95% CI: 0.68-0.73).

Based on our observations from disrupting the spectral and temporal patterns, we hypothesized that the predictions of our model are guided more by the spectral composition of the measurements than by their temporal patterns. Hence, we sought to predict preterm births using shorter EHG recordings. The duration of EHG recordings, usually between 30 and 60 minutes, is an important hindrance to their implementation in clinical settings, where personnel resources are often limited [23, 44].

To test this hypothesis, we trained and tested our model using cropped STFTs, as shown in Fig. 3e. We removed columns at the beginning and at the end of the STFTs to simulate shorter EHG measurements. Since the initial point selected for these shortened STFTs slightly affects the resulting AUC, we selected a random initial point for each shortened sample. Remarkably, the performance of our model decreased only marginally with decreasing measurement duration, as shown in Fig. 3f. When we trained our model using one-minute long recordings, we could predict preterm births with an AUC of 0.73 (95% CI: 0.71-0.75), which is only slightly lower than the 0.75 (95% CI: 0.74-0.77) AUC we obtained using the entire 30-minute long recordings.

## Discussion

We developed a deep learning method to predict preterm births from EHG measurements and clinical information obtained from two public databases. We predicted preterm births with good accuracy directly from the data and without using handcrafted features, manual annotations, or simultaneous tocography measurements. Thus, our method potentially enables automatic prediction of preterm births from EHG recordings.

To assess the performance of our method from the perspective of clinical practice, we compared the performance of our method with other technologies and methods to predict preterm births, as shown in Table 3. For this comparison, we included results only from studies published in peer-reviewed journals, with sound methodology, that reported the AUC of the predictions, and which included at least 50 pregnant mothers. Similarly to the datasets used in this work, the results reported in these studies correspond to obstetric populations excluding medically induced births. From this comparison, we observe that the performance of our method is superior to the performance of existing methods to predict preterm births that take place before 37 complete weeks of gestation. Importantly, our method outperforms the gold standard biomarkers of preterm birth, i.e., cervical length and fibronectin alpha, in this task. Moreover, the performance of our method in predicting preterm births in asymptomatic mothers around their 31st week of gestation is relatively close to the performance of the gold standard tests in predicting preterm birth within only one week in mothers with symptoms of preterm labor. Our results support previous findings suggesting that preterm birth can be predicted in asymptomatic mothers by using EHG measurements from around the 31st week of gestation [19, 45].

**Table 3:** Accuracy of several technologies and methods to predict preterm births.

| Technology | Population | AUC |
| --- | --- | --- |
| Prediction of birth before 37 weeks of gestation |  |  |
| Clinical information [46] | Singleton pregnancies, asymptomatic mothers | 0.64 (0.61-0.67) |
| Our method (clinical information alone) | Asymptomatic mothers | 0.56 (0.54-0.59) |
| Fibronectin alpha [9] | Asymptomatic mothers | 0.65 (0.63-0.66) |
| Fibronectin alpha [9] | Symptomatic mothers | 0.71 (0.69-0.73) |
| Cervical length [12] | Nulliparous, singleton pregnancies, asymptomatic mothers | 0.67 (0.64-0.70) |
| Clinical information and placental measurements [46] | Singleton pregnancies, asymptomatic mothers | 0.72 (0.66-0.77) |
| EHG [24] | Asymptomatic mothers | 0.60 |
| EHG [22, 45]<sup>a</sup> | Asymptomatic mothers | 0.61 |
| EHG [19, 22]<sup>a</sup> | Asymptomatic mothers | 0.65 |
| EHG [28]<sup>b</sup> | Asymptomatic mothers | 0.84 |
| EHG [27]<sup>b</sup> | Asymptomatic mothers | 0.97 |
| EHG [29] | Asymptomatic mothers | 0.69 |
| Our method (EHG alone) | Asymptomatic mothers | 0.75 (0.74-0.77) |
| Our method (combined classification model) | Asymptomatic mothers | 0.80 (0.79-0.82) |
| Prediction of preterm birth within one week |  |  |
| Fibronectin alpha [9] | Symptomatic mothers | 0.84 (0.80-0.87) |
| Cervical length [10] | Symptomatic mothers | 0.84<sup>c</sup> |

<sup>a</sup> We considered the performance of these methods after their methodology was corrected by Vandewiele et al. [22]. <sup>b</sup> Performance was evaluated on a combination of real and synthetic data. The performance on real data alone is not reported. <sup>c</sup> The AUC was obtained by extrapolating the ROC curve. AUCs are presented with 95% CI in parenthesis when available.

Additionally, we investigated how the temporal and spectral components of the EHG measurements contribute to our model’s predictions. We observed that the higher frequency components of the EHG measurements are more predictive of preterm births. A possible explanation for this phenomenon is that the higher frequency bands contain spectral harmonics of the electrical activity in EHG measurements and more spectral information may be coded in the higher frequency bands. However, further research is needed to decipher the sources of the various spectral components of EHG measurements.

Importantly, we observed that the temporal patterns measured in EHG measurements are not crucial to predicting preterm births. This observation agrees with the results published by Iams et al., who showed that the frequency of uterine contractions is not predictive of preterm births [47]. Moreover, this observation also might explain the inability of tocography, which measures the temporal patterns of uterine contractions, to predict preterm births [13]. Inspired by this observation, and specifically by our results presented in Fig. 3d, we explored whether we could use shorter EHG measurements to predict preterm births.

Notably, we found that shortening the EHG measurements did not substantially degrade the performance of our model in predicting preterm births. Our findings using short EHG measurements, together with the positive results obtained by Jager et al. for predicting preterm birth using EHG and tocodynamometer measurements from only short contractile or non-contractile intervals, suggest that shorter EHG recordings could be sufficient to predict preterm births [19]. From the perspective of clinical adoption, a shorter recording is easier for the mother and saves cost. Moreover, the shortened recording time combined with the automaticity of our method facilitates at-home implementations.

Whereas the classification and regression models could predict preterm births with good accuracy, surprisingly, the regression models could not predict the gestational ages at delivery accurately, as shown in Supplementary Fig. 3. This effect can be explained by the pathology of preterm births and by analyzing the distribution of the gestational ages at delivery. Preterm birth is an abnormal physiological condition, not just a pregnancy that happened to end early. Therefore, we can expect that physiological measurements, such as EHG recordings, may show a stronger dichotomy between pregnancies that end with either preterm or term deliveries than is shown in continuous characteristics correlated with gestational age at delivery.

We observe the dichotomous aspect of preterm and term births through the distribution of the gestational ages at delivery, shown in Supplementary Fig. 1 and in Supplementary Fig. 2. The distribution of the gestational ages at birth of the mothers included in this work only from the TPEHG DB is left-skewed and does not appear to follow a Gaussian distribution, as shown in Supplementary Fig. 2d. This skewness may can be caused by either an excess of preterm births compared to what would be expected if the gestational ages at birth followed a Gaussian distribution and by the induction of postterm births, which can skew the distribution towards earlier deliveries. However, when we exclude the preterm births the distribution of gestational ages at birth appears to follow a Gaussian distribution, as shown in Supplementary Fig. 2h. This observation suggests that the skewness results from an over-representation of preterm births rather than from the induction of postterm births. Since the gestational ages at delivery do not follow a Gaussian distribution where the left tail accounts for preterm births, we suggest that the dynamics that dictate the gestational age at delivery do not follow a continuum between preterm and term births. Therefore, we propose that predicting the gestational age at delivery is more complicated than predicting preterm births using categorical outputs.

The significance of predicting preterm births in mothers without symptoms of preterm labor and several weeks before delivery is that it can be helpful in delaying preterm births and improving their outcomes. For example, clinical providers can prescribe progesterone to these mothers to prolong their pregnancies [48, 49]. Additionally, medical providers could more frequently screen mothers at high risk of preterm birth to identify and treat hypertensive disease and cervical insufficiency [50, 51]. Moreover, anticipating preterm births can be useful in planning for the birth to take place at a hospital with a neonatal intensive care unit (NICU), rather than at home, in birthing centers, or in hospitals without a NICU, thus avoiding ambulance transport and admission delays and improving outcomes [52, 53, 54]. Furthermore, identifying asymptomatic mothers at high risk of preterm birth may help researchers assess the efficacy of potential approaches and therapies to delay preterm births and improve their outcomes.

Although machine learning algorithms can contribute to improving healthcare and much research is yielding advances in this field, important challenges remain [55, 56]. For example, machine learning predictions usually lack interpretability, meaning that it is challenging to identify the causes justifying the algorithms’ predictions [55, 56]. In our case, although our predictions could influence pregnancy management, our predictions would need to be supplemented with additional medical examinations to determine which therapies are more likely to reduce the risk of preterm birth and improve its outcomes. Additionally, machine learning algorithms in healthcare settings need to be carefully developed to protect data privacy and to prevent social biases from driving the predictions [56, 57, 58].

Despite the limitations of machine learning algorithms for developing medical devices, the number of medical products based on machine learning is steadily increasing thanks to their good performance [59]. By predicting preterm births with good accuracy directly from the measurements, while avoiding data leakage, our work is a step forward towards developing a medical device for predicting preterm births from EHG measurements using deep learning.

Our work is limited by the etiology of preterm birth and the dataset that we used to develop our models. Because preterm birth is a syndrome with many causes, it is most likely that no single physiological measurement will predict preterm births with perfect or nearly perfect accuracy [5, 60]. A combination of measurements of various physiological processes is likely to produce better results [5, 61].

The limited size of the datasets employed limits our work. We evaluated our prediction models using cross-validation rather than separating a subset of the data exclusively for testing after developing our models, because such a testing set would be too small for accurate performance evaluation [39]. For example, if we set apart 20% of the data for the final testing, this dataset would contain five preterm and 25 term samples. Moreover, all the samples in the dataset were acquired in a single hospital, and thus our model may not generalize well to measurements from mothers in different populations. Additionally, the datasets used in our work, and in those mentioned in Table 3, excluded medically induced births, and therefore, these populations may differ from general obstetric populations. Interestingly, Erkamp et al. found similar screening performance for preterm birth using sonographic measurements when either including or excluding medically induced births from their analysis [46]. A larger database, preferably acquired across multiple healthcare centers, could rectify these limitations. Specifically, a larger database would enable us to separate a subset of samples to further evaluate the generalizability of our model. Moreover, because of the limited size of the database, we trained a small neural network with a limited number of parameters. In the future, a larger database would also enable us to train larger and more complex prediction models for better results [62].

Our work can be expanded to improve its performance and clinical value. First, following the same approach we used to combine EHG measurements with clinical data to predict preterm births, our method could incorporate other data, such as cervical length and fibronectin alpha measurements, which are likely to improve its performance. Additionally, to track the evolution of EHG activity towards birth and develop a dynamic prediction model, multiple EHG measurements could be recorded throughout pregnancy for each mother. Moreover, EHG measurements could be recorded from mothers presenting symptoms of preterm labor to develop a deep learning model for predicting preterm birth within one week. Lastly, our work could be integrated with models connecting surface EHGs with uterine sources to include anatomical and physiological information for making predictions [63, 64].

### Conclusions

In summary, we developed a deep learning model to predict preterm births using clinical information and EHG measurements. Our method predicted preterm births in pregnant mothers without symptoms of preterm labor more accurately than existing technologies. We also showed that preterm births can be predicted using short EHG recordings. Our work and results are useful for developing applications to predict preterm births early during pregnancy and for ultimately improving their outcomes.

## Data Availability

All data produced in the present work are contained in the manuscript.

## Acknowledgement

This research was supported by the McDonnell International Scholars Academy at Washington University in St. Louis.

