## Supplementary materials for "Predicting preterm births from electrohysterogram recordings via deep learning"


### Supplementary information

#### Supplementary Fig. 1

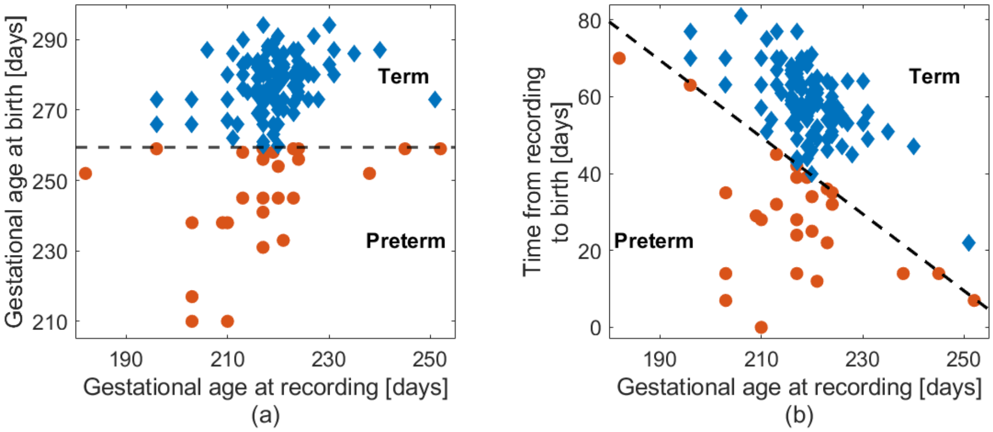


Supplementary Fig. 1: Distribution of gestational ages at the times of recording and at the times of birth. (a) Gestational ages at birth plotted against gestational ages at the times of recording. (b) Elapsed times between recordings and births, plotted against gestational ages at recording. (a), (b) The dashed black lines separate preterm (red circles) and term births (blue diamonds).

### Supplementary Fig. 2

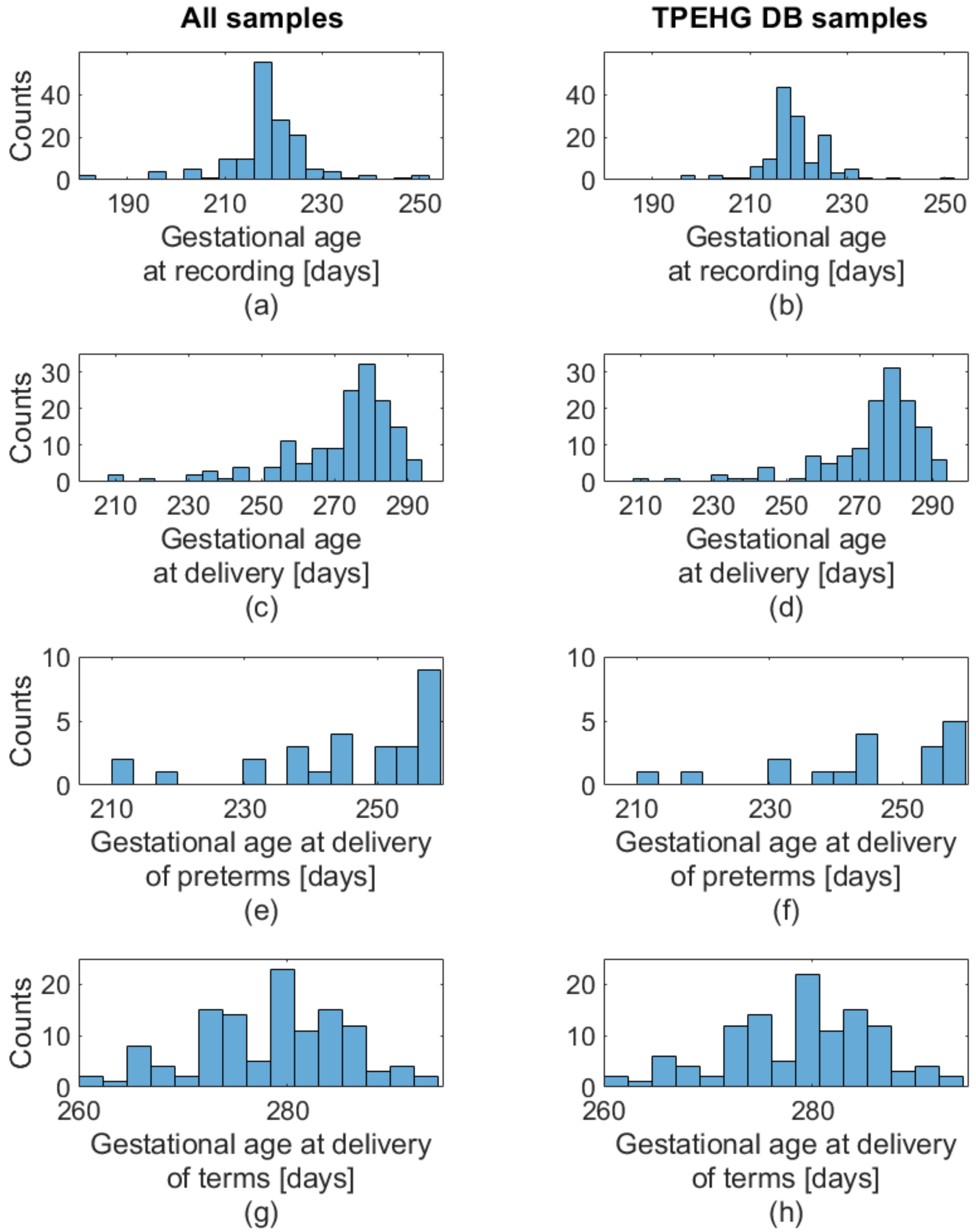


Supplementary Fig. 2: Distribution of gestational ages at the times of recording and delivery. Note dissimilar scale ranges between graph pairs. (a) Histogram of the gestational ages when the EHGs were recorded. (b) Same as **a**, but using only the samples used from the TPEHG DB. (c) Histogram of the gestational ages at the times of delivery. This distribution is left skewed (skewness = -1.7) and does not appear to follow a Gaussian distribution ($p = 7.7 \times 10^{-10}$). (d) Same as (c), but using only the samples used from the TPEHG DB. This distribution is also left skewed (skewness = -1.9) and does not appear to follow a Gaussian distribution ($p = 6.4 \times 10^{-10}$). (e) Histogram of the gestational ages at the times of delivery for the preterm births. Preterm births are more common at older gestational ages. (f) Same as (e), but using only the samples used from the TPEHG DB. (g) Histogram of the gestational ages at delivery for the term births. This distribution appears to follow a Gaussian distribution ($p = 0.06$). (h) Same as (g), but using only the samples used from the TPEHG DB. This distribution also appears to follow a Gaussian distribution ($p = 0.07$). (c), (d), (g), (h), Normality was assessed using the Shapiro-Wilk test.

### Supplementary Fig. 3

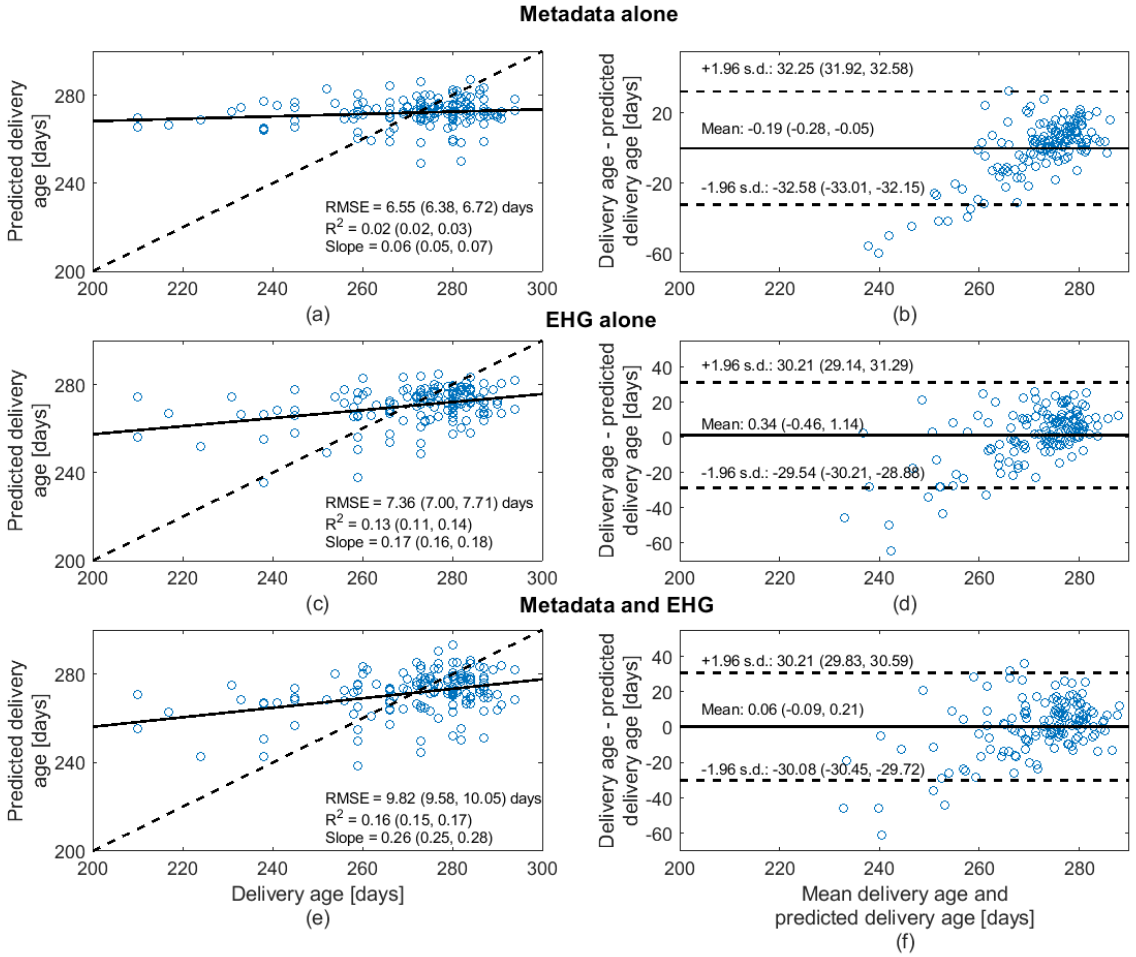


Supplementary Fig. 3: Predictions of gestational ages at delivery using the regression models. (a) Predicted gestational ages at birth, using the clinical information alone plotted against the true gestational ages at birth. Each blue circle shows the gestational age at delivery, predicted based on the clinical information and the true gestational age at delivery for a single mother. The solid black line represents the linear fit between the predictions and the true values. The dashed black line represents a perfect correspondence between predictions and true values. The legend shows the root mean square error (RMSE) of the predictions, the coefficient of determination ($R^2$) of the predictions, and the slope of the linear fit. (b) Bland–Altman plot for the predicted gestational ages at birth, using the clinical information alone and the true gestational ages at birth. Each blue circle represents the difference between predicted and true gestational ages at birth, and the mean of these values. The solid and dashed black lines show the mean of the difference between the predicted and the true values, and the 95% limits of agreement, calculated as mean $\pm$ 1.96 standard deviations, respectively. (c) Similar to (a), but using the predictions based on EHG measurements alone. (d) Similar to (b), but using the predictions based on EHG measurements alone. (e) Similar to (a), but using the predictions based on clinical information combined with EHG measurements. (f) Similar to (b), but using the predictions based on clinical information combined with EHG measurements. All values are presented as mean with 95% CI.
